# Non-contact fingertip video to differentiate dengue haemorrhagic fever in plasma leakage from dengue fever: A prospective proof-of-concept cohort study

**DOI:** 10.64898/2026.10.30.26366674

**Authors:** M. Thisara S. Perera, Shanali Thathsarani Manamperi, S.A.A. Janindu M. Athukorala, K.W. Sameera M. Wijayawardhana, Akhila Rakshitha Wimalasundera, Salomi Arasaratnam, D.M. Janitha Prathapa, Yudara Kularathne

## Abstract

Most patients with dengue recover without intervention, but a minority develop plasma leakage that threatens the circulation, and differentiating them is the central problem in dengue care. Platelet count is the marker in routine serial use and is itself one of the four defining criteria of dengue haemorrhagic fever (DHF), so thrombocytopenia in dengue fever (DF) can overlap with DHF on the very marker used to distinguish them. We asked whether a non-contact optical signal carries discriminative information beyond platelet count.

We conducted a prospective observational cohort study at a national dengue referral centre. All DHF recordings were obtained at or after plasma-leakage onset, so the analysis compares established clinical states rather than pre-leakage risk. Serial non-contact fingertip videos were recorded with contemporaneous clinical and haematological measurements. In a held-out set of 50 recordings (20 DHF, 30 DF), video-derived probability separated the groups with an area under the receiver operating characteristic curve (AUC) of 0.963 (95% CI 0.918–1.000; 0.957 averaged across eight architectures). In the 49 recordings with a platelet count, the video AUC was 0.961 against 0.888 for platelet count (difference 0.074, p = 0.19). In exploratory analyses the video added information to platelet count (likelihood-ratio p < 0.001), showed no significant within-group correlation with platelet count, and retained an AUC of 0.970 against thrombocytopenic DF, where platelet count fell to 0.759. Bedside pulse rate, body temperature and haematocrit did not separate the groups and were unrelated to the video score.

A ∼30-second fingertip video separated DHF in plasma leakage from DF in this test set, and exploratory analyses suggest it carries information beyond platelet count. These findings are proof of concept from single-centre internal validation, not evidence of pre-leakage prediction; prospective pre-leakage studies and external validation are needed to establish whether the signal can support earlier risk stratification.

**Author summary:** Dengue is common in Sri Lanka. While most patients recover, a subset experiences a critical leakage of fluid from their blood vessels, which can progress to shock, bleeding and organ dysfunction if untreated. We monitor platelet counts because they track disease progression, but counts can fall in patients who leak fluid and in those who do not, so a low count alone cannot separate the two groups. We investigated whether a brief, non-contact fingertip video could tell these groups apart, beyond platelet count. We recorded dengue haemorrhagic fever videos at or after the onset of leakage, and in test recordings held out from training, the video separated the two groups and, in exploratory analyses, appeared to add information beyond the platelet count. This held among patients with low platelet counts in both groups. This single-hospital study does not establish a diagnostic test for clinical use. Its significance is in showing that a simple optical recording can detect a difference associated with established disease. Our next question is whether this difference can be detected before leakage begins; if so, the same recording could eventually help identify, at a population level, which dengue patients are most at risk of these complications.

## 1. Introduction

### 1.1 The clinical problem

Dengue is the most widespread arboviral infection of humans, with an estimated 390 million infections annually, of which roughly a quarter are clinically apparent [1]. In Sri Lanka the disease is hyperendemic, with recurrent large epidemics and a hospital burden that periodically exceeds the capacity of the acute care system [2]. The great majority of these patients have an illness that will resolve without intervention. A minority develop plasma leakage severe enough to threaten the circulation, and in that minority the difference between timely and delayed fluid resuscitation is a difference in survival.

The clinical task is therefore not diagnosis of dengue, which is usually straightforward, but discrimination within dengue, early enough to act on. In practice this is attempted through serial full blood counts and observation for the WHO warning signs.

Several approaches have been proposed to anticipate severe dengue before it becomes clinically obvious. Host genetic and gene-expression studies have linked common variants and inflammatory signatures to dengue shock syndrome and severity [3,4], viraemia kinetics and platelet trajectories have been modelled directly [5,6], and an eight-gene machine-learning classifier built on these signals improved prediction of severe progression [7]. In parallel, wearable contact photoplethysmography (PPG) has been explored in hospitalised dengue patients: an artificial-intelligence approach applied to 24-hour wearable PPG identified volume-status changes consistent with shock [8], a multimodal transformer applied to continuous PPG and basic clinical data forecast clinical state 2 hours ahead [9], and a convolutional neural network classifier classified adults into three severity classes from wearable PPG waveforms (precision 0.79, recall 0.77) and distinguished acute from convalescent illness in children (precision and recall 0.89) [10]. Closer to the present approach, smartphone-camera fingertip video has itself been used to estimate platelet count by regression (R² = 0.88) [11] and to classify dengue-positive from dengue-negative individuals with 88% accuracy [12], both using contact fingertip video — the finger pressed against the camera and flash — rather than the non-contact, at-a-distance remote photoplethysmography (rPPG) used here. None of these approaches has asked whether an optical signal separates dengue haemorrhagic fever from dengue fever independently of platelet count once thrombocytopenia is already present in both groups, the question this study addresses.

No differences in dengue incidence or outcome between sociodemographic groups in this population are established; the cohort is drawn from patients admitted to a single national referral centre.

### 1.2 The platelet count: the reference marker and its limits

A rising haematocrit, the ultrasonographic signs of leakage and the WHO warning signs share a structural limitation: they are defined in terms of the event they are meant to anticipate, and each detects plasma leakage only once it is under way [13,14]. The platelet count is different, and it is the marker this study uses as its reference, for two reasons. First, it is the quantity in routine serial use: a falling count is tracked daily in every dengue ward, and its discriminative value, although greatest late in the illness, is real. Second, and more importantly here, thrombocytopenia is not merely associated with dengue haemorrhagic fever — it is one of the four defining criteria of the 1997 WHO case definition [15]. This has two consequences that shape the analysis. Because the platelet count is built into the label itself, it will correlate with the diagnosis by construction; a marker that exceeds it therefore does so conservatively, against a comparator that has been handed part of the answer. Since dengue fever can produce thrombocytopenia without the remaining criteria being met, the two forms genuinely overlap on the marker. That overlap is precisely where a better marker must prove itself. The useful question is therefore not whether a new marker can predict haemorrhagic fever — a question the case definition makes partly circular — but whether a new marker carries discriminative information the platelet count does not.

This study is not an exception to the timing limitation above: because every DHF recording in the study cohort was obtained at or after the onset of leakage (Section 2.5), the video signal is evaluated in the same post-onset window as the platelet count, not a window the platelet count already occupies alone. What differs is the comparator, not the window. Showing that the signal carries information beyond the platelet count in that window is the necessary first step before the harder, and currently unanswered, question of whether it can be detected earlier (Discussion).

### 1.3 A candidate signal: optical measurement of the peripheral microcirculation

Three physiological chains connect the plasma-leakage lesion of dengue haemorrhagic fever to a signal recorded at the fingertip, and they are not mutually exclusive.

The first is endothelial. Dengue haemorrhagic fever is associated with degradation of the endothelial glycocalyx and of the tight-and adherens-junction proteins that seal the vascular wall, and the degree of degradation tracks the severity of plasma leakage: elevated syndecan-1 and chondroitin sulfate were associated with plasma leakage, and syndecan-1 and claudin-5 with severe leakage, in a prospective Indonesian cohort [16], and heparanase-mediated glycocalyx shedding correlates with plasma leakage markers including ascites and gallbladder wall thickening in a second cohort [17]. A mast-cell-derived tryptase that opens endothelial tight junctions is sufficient to reproduce plasma loss and hypovolaemic shock experimentally, and plasma tryptase levels track DHF severity grade in two independent human cohorts [18]. Direct microcirculatory imaging shows the same lesion functionally: the sublingual proportion of perfused vessels and microvascular flow index are lower in patients with leakage, correlating with circulating markers of endothelial activation [19]. This is the lesion the study sets out to detect — but its established outputs (circulating protein markers, sublingual imaging) are not what a fingertip video records.

The second is haemodynamic. Plasma loss reduces effective circulating volume, and the earliest compensation for reduced central volume is autonomically mediated redistribution of blood flow away from the skin and toward the vital organs — a response that is graded to the degree of volume loss and measurable through its effect on peripheral vascular tone before conventional vital signs change [20]. This compensatory reserve response, modelled originally under simulated hypovolaemia, has been shown to transfer to true haemorrhage: compensatory reserve tracked equivalently between simulated central hypovolaemia and graded blood loss in non-human primates [21], and among candidate bedside measures, an arterial-waveform-derived reserve index discriminated the onset of decompensation better than pulse pressure variability or a simple perfusion index [22]. Because this redistribution acts on the cutaneous and acral circulation specifically, and precedes overt haemodynamic decompensation, it offers a route by which plasma leakage could alter a fingertip signal without any local change in vessel permeability at the recording site itself.

The third is optical. What a camera-based photoplethysmographic signal records is not permeability or perfusion directly, but the small cyclical change in optical path length through the fingertip skin produced by each heartbeat’s arrival — a signal jointly determined by stroke volume, arterial and arteriolar compliance, and local vasomotor tone, not by vessel wall permeability [23,24]. Either of the first two chains — a change in local vascular tone from compensatory vasoconstriction, or a change in the pulse waveform from altered circulating volume — is capable of altering this optical signal without the glycocalyx lesion itself being optically visible at the fingertip.

These three chains are not exclusive, and this study cannot adjudicate between them: the video model is trained and evaluated as a whole, with no feature-attribution or channel-removal analysis reported (Section 4). What they establish is that a plausible physiological route exists from the leakage lesion to a peripherally recorded optical signal by at least three non-exclusive mechanisms — the warrant for testing the signal at all, not evidence for which mechanism operates.

None of these three chains is a claim that fingertip rPPG measures microvascular permeability. Permeability, perfusion and pulsatile blood volume are distinct physiological quantities: a vessel can be leaky with no change in pulsatile amplitude, and pulsatile amplitude varies with vasomotor tone, stroke volume and temperature independently of permeability. What the three mechanisms above establish is only that a leakage state is plausibly accompanied by a change in one or more of the quantities that do determine the optical signal — local vasomotor tone and the arterial pulse waveform — not that the optical signal is a direct readout of the lesion itself.

Remote photoplethysmography derived from ordinary video samples a cutaneous microvascular bed directly, requires no contact, no consumable and no venepuncture, and can be repeated as often as the clinical question demands [25,26]. We record at the fingertip because peripheral perfusion is under regional autonomic control, and abnormal peripheral perfusion at the extremities carries prognostic information in critically ill patients [27]. In healthy volunteers subjected to graded central hypovolaemia, the peripheral pulse-oximetry perfusion signal fell by about 40% at the mildest step, while stroke volume, heart rate and blood pressure had changed little [28]. Video photoplethysmography is sensitive to subject motion and illumination [29], and potentially to skin pigmentation and camera behaviour; the acquisition and analysis were designed around that objection (Sections 2.5–2.7).

### 1.4 The hypothesis and four expectations

The hypothesis is comparative, and it decomposes into four testable expectations. The video signal should (i) separate the two forms at least as well as the platelet count; (ii) add information to the platelet count; (iii) retain its separation when DF is restricted to patients with platelet counts in the DHF range — the decisive test, because there the reference marker has been largely deprived of its built-in advantage; and (iv) be uncorrelated with the platelet count within each group, so that any separation it achieves cannot be a restatement of the count.

### 1.5 What would refute it

The hypothesis would be weakened if video probability tracked platelet count within either group, if separation disappeared among patients with overlapping platelet counts, or if the result could be explained by bedside pulse rate, body temperature or recording conditions. Sections 3.2 and 3.3 report the comparisons and sensitivity analyses available in the study cohort. These analyses test potential alternative explanations within the available data; they do not establish a physiological mechanism or exclude all residual confounding.

### 1.6 Objectives

The primary objective was to determine whether a signal derived from short fingertip videos separates the DHF-in-plasma-leakage state from the DF state, with information beyond platelet count, as specified in Section 1.4. Demonstrating additional separation after leakage is a proof-of-concept step: it establishes a candidate optical signal for the more clinically important question of whether a difference can be detected before leakage. The study does not evaluate a triage instrument or establish benefit for clinical decision-making.

## 2. Materials and methods

### 2.1 Study design and setting

Prospective observational cohort study conducted at the National Institute of Infectious Diseases, Angoda, Sri Lanka, the national referral centre for dengue and the site of the WHO Collaborating Centre for Dengue Clinical Management and Clinical Research. Patients were recruited between May 2026 and August 2026.

Ethical approval for this study was obtained from the Ethics Review Committee of the National Institute of Infectious Diseases (NIID), Sri Lanka (ERC Reference No. P/04/02/2026). Written informed consent was obtained prior to enrolment from all adult participants and, for participants below 18 years of age, from a parent or guardian together with the participant’s assent. To protect participant privacy, all research data were pseudonymised using unique study identification numbers, and personally identifiable information was not included in the research dataset. Video recordings were limited to the participant’s finger. Clinical data and video recordings were treated as confidential research data and stored securely, with video files maintained in a private Amazon S3 bucket with public access disabled, encryption at rest, and secure HTTPS/TLS transmission. Access to the data was restricted to authorised members of the research team through controlled permissions. The study was conducted in accordance with the principles of the Declaration of Helsinki. Patients and the public were not involved in the design, conduct, reporting or dissemination of this study.

### 2.2 Participants

Eligible patients were adults and adolescents (>14 years) presenting with an acute febrile illness clinically consistent with dengue and subsequently confirmed by laboratory testing. Dengue diagnosis was confirmed by either NS1 antigen or positive IgM antibodies. A convenience sampling method was used. No formal sample size calculation was performed. All eligible patients recorded during the study period were included, and the resulting size was fixed by recruitment rather than chosen; the study is presented as a proof of concept.

### 2.3 Reference standard and case definition

The outcome was the final clinical diagnosis of DF or DHF assigned according to WHO 1997/SEARO 2011 criteria [15,30], applied to the complete admission record rather than to the state of the patient at the time of any individual recording. The 1997 criteria were used in preference to the 2009 classification because the hypothesis concerns plasma leakage specifically, and the DHF category encodes that pathophysiology in a way that the 2009 severity-based classification deliberately does not [31].

Diagnosis was assigned in two steps. First, if the patient met DHF criteria at any point during the admission, the final diagnosis was DHF. Second, patients coded as DF at a given visit were re-examined during the hospital stay and recoded as DHF if they later met DHF criteria. DHF was confirmed when there was ultrasonographic evidence of leakage. All patients admitted to the study centre undergo twice-daily bedside scans as routine practice to detect leakage objectively. Patients whose labels remained inconsistent across visits were removed before the cohort was frozen (Section 3.1).

The final diagnosis was assigned by middle-grade doctors of the attending team who were trained in dengue bedside ultrasound scanning, and bedside ultrasound was performed by the same cadre. Diagnosis was assigned during routine clinical care by clinicians who had no access to the video recordings or to any model output. Video recording was performed by a member of the study team who was not involved in patient management or diagnostic decision-making, trained to acquire the recording protocol (Section 2.5) in consultation with the engineering team.

Every recording was labelled with the day of illness, counted from the patient-reported onset of fever, with the first day of fever designated day 1. All analyses are indexed on this variable rather than on calendar date.

Recordings were classified by final diagnosis and clinical state at the time of recording using the five-state framework in Table 1. The framework is retained to distinguish the observed states and to define strata for separate pre-leakage and phase-specific analyses. It does not imply that those analyses were performed in the study cohort.

**Table 1.**
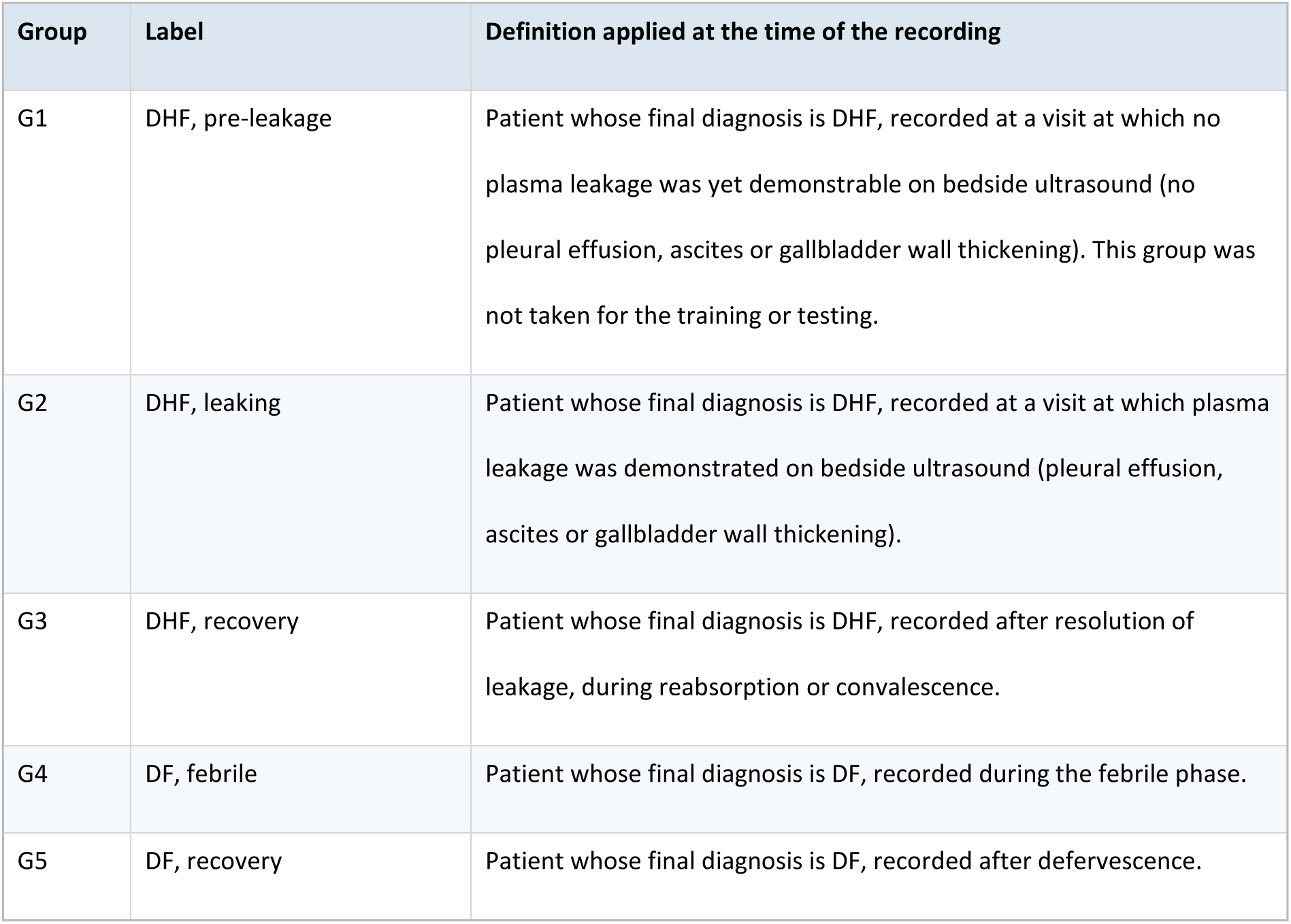
Clinical state framework by final diagnosis and state at recording. G1 defines the pre-leakage DHF stratum for separate analyses. These definitions do not establish phase-invariance or imply that all five strata contributed to the present model.

The present comparison pools the available post-onset DHF recordings, including recovery, against DF.

### 2.4 Reference measurements and ground truth

At each study visit the following were recorded contemporaneously with the video.

- Full blood count including platelet count, haematocrit and white cell count, with the serial values from the admission used to establish the haematocrit trajectory and the platelet nadir.
- Vital signs including temperature, pulse rate, blood pressure, respiratory rate and capillary refill time.
- Ultrasonographic assessment for pleural effusion, ascites and gallbladder wall thickening as the ground truth of leaking evidence.

### 2.5 Video acquisition — clinical protocol

Recordings were obtained from the fingertip. Patients were lying or semi-recumbent and rested for at least 5 minutes before recording, in a room with controlled lighting, with the hand supported to minimise movement.

All recordings were made with the same smartphone (iPhone 14 Pro Max, iOS 26.x, updated during the study), in video mode, under the same recording conditions throughout the study. The video files are recorded at 30 frames per second (median measured rate 29.96), in portrait orientation, and last 22–31 s. The patient pointed the index finger with the other fingers flexed, and the camera filmed the finger from in front, without contact (Fig 1A).

**Fig 1.** How a probability is obtained from a fingertip video. (A) The fingertip region located by the segmentation network (green) on one frame of six held-out test recordings (three DF, three DHF), showing the non-contact recording geometry and the variation in background and skin tone. (B) The six processing steps. The same code is used during training, evaluation and in the deployed software; the networks receive the nine-channel signal, not the image.

Even with one device and one protocol, small differences between recordings in lighting and in how large the finger appears in the frame can be seen; this is addressed by checking recording conditions before modelling and by normalising each signal to its own baseline within the analysis (Section 2.6, S1 Text).

### 2.6 Signal extraction and feature derivation

The fingertip is located automatically on every frame by an instance-segmentation model [32], and the colour of the fingertip skin is tracked over time and expressed relative to its own recent baseline, reducing differences in overall brightness and skin tone between recordings (skin tone was not measured). Established remote photoplethysmography methods (plane-orthogonal-to-skin [23] and chrominance [24]) turn this colour signal into nine channels — waveforms in the cardiac frequency band and a slow band capturing breathing and vasomotor oscillation — none of which is a heart-rate value; the channels are cut into 10-second windows before being given to the model. No clinical, laboratory, demographic or timing variable enters the model. The full processing specification, including all filter settings, frequency bands and parameter values, is given in S1 Text, and S2 Fig illustrates the signal construction for one recording.

A signal-quality rule, based on how much of the recording is usable, was applied to every recording before modelling; every held-out test recording passed it (S1 Text). If the fingertip cannot be located or a recording fails this rule, no probability is issued and the recording is repeated; the model is not intended to return a result from a recording it cannot process. Signal quality was not compared across sociodemographic groups.

### 2.7 Model development and validation

The final model is an ensemble of five small convolutional neural networks of the DeepPhys type [33], adapted from video frames to the nine-channel signal described in Section 2.6; the network reads the physiological signal rather than the image, which restricts its input to colour fluctuation over time and is intended to reduce its sensitivity to background and to the absolute colour of the skin. No clinical, laboratory, demographic or timing variable enters the model. Eight architectures from four families (convolutional, residual, recurrent and attention-based) were trained under an identical protocol, and the final architecture was chosen by the pre-specified rule described below; the comparison is reported in Section 3.2 and S1 Fig. The exact network design, parameter count and training protocol are given in S1 Text.

#### Inputs and outputs

The output is one probability of DHF per recording, obtained by averaging the probabilities across the recording’s 10-second windows (median 7 per recording) and the five networks. A recording is classified as DHF if the probability is at least 0.50; this threshold was fixed before the test set was scored and was not tuned. DHF is the positive class throughout.

To avoid training a model dominated by one class, the DF class was reduced to a size comparable with the DHF class. All DHF recordings were retained, and their mean fingertip area in the frame served as the reference for finger-to-camera distance; the DF recordings whose fingertip area lay closest to that reference were selected, so that the two classes had comparable size and recording geometry (Fig 2). Separately, all recordings were checked for comparable recording conditions (lighting and fingertip size in the frame) to confirm that they had been acquired under the same protocol. Selection was completed before any model was trained, without reference to any model output. These were divided into a training partition and a held-out test partition (Fig 2). The partition was made at the level of the patient: all recordings from the same patient were placed in the same partition, and no patient contributed recordings to both. This was verified against the cohort workbook, in which every recording is assigned to its partition and every test recording is matched to a single row by hospital number, day of fever and video file name. The inner folds used within the training partition, only to choose each network’s stopping epoch, never contain test recordings and cannot influence the test result.

**Fig 2.** Selection and partition of the finger-video dataset. The cohort comprised 311 recordings from 267 patients (DF, 249 recordings from 211 patients; DHF, 62 recordings from 56 patients). All 62 DHF recordings were retained, and their mean fingertip area in the frame served as the reference for finger-to-camera distance. From the 249 DF recordings, the 88 whose fingertip area lay closest to that reference were selected, giving a modelling dataset of 150 recordings (62 DHF, 88 DF) from 136 patients with comparable class sizes and comparable recording geometry. Selection was completed before any model was trained and used no signal, label or outcome information beyond the recording’s class. The modelling dataset was split by patient into a training partition (42 DHF recordings from 36 patients; 58 DF recordings from 51 patients) and a held-out test partition (20 DHF recordings from 20 patients; 30 DF recordings from 29 patients); all recordings from a patient were assigned to the same partition, and no patient appears in both.

The model was not given the phase or the day of fever, and no phase-specific model was fitted. The distribution of DHF recordings by clinical state is reported in Sections 3.1 and 3.2.

The test set was not used for training, standardisation, epoch selection or early stopping. It was used once, to rank the eight candidate architectures, and the architecture with the highest test balanced accuracy was retained. To quantify the effect of this selection, the test set was also used to evaluate the unweighted average of all eight architectures, which involves no selection.

Proportions are reported with Wilson 95% intervals [34]. The area under the receiver operating characteristic curve (AUC) is reported with DeLong 95% intervals, and paired AUCs on the same recordings are compared with the DeLong test [35]; a patient-clustered bootstrap (2,000 resamples) and a label-permutation test (10,000 permutations) were used as checks. Calibration was assessed by the calibration intercept and slope from logistic recalibration, the Brier score and observed against predicted risk by quintile [36]. Discrimination was also summarised by the average precision (area under the precision–recall curve), which is informative when the classes differ in size. As a post-hoc check that the single patient with two held-out recordings did not influence the result, the held-out AUC was recomputed using one recording per patient. The contribution of the video beyond platelet count was assessed on the test recordings with logistic models containing the log-odds of the video probability, log platelet count, age and sex, using likelihood-ratio tests and leave-one-out cross-validated AUC with ridge-penalised coefficients (penalty C = 1 on standardised predictors). All secondary analyses (comparison with platelet count, the thrombocytopenic subgroup, correlations and sensitivity analyses) were performed on the same held-out test set after the model was fixed; they did not alter the model, were not corrected for multiple comparisons, and are exploratory. The held-out AUC was the primary outcome. Reporting follows TRIPOD+AI [37]; the completed checklist is provided as S1 Checklist.

No fairness-specific method was applied; as a post-hoc check, discrimination was examined by sex and by age band (below, or at or above, the test-set median of 30 years), with subgroup AUCs compared by a two-sided z-test on their DeLong standard errors; the video probability was related to age (Spearman) and compared between sexes (Mann–Whitney) (Section 3.3).

Recordings were excluded only for the reasons given in Section 3.1 and Fig 2. Where a clinical variable was missing for a recording, that recording was omitted from the analysis of that variable alone, and the number of recordings is reported with each analysis.

### 2.8 Statistical analysis of clinical variables

Clinical and haematological variables are summarised as mean with standard deviation or median with interquartile range according to distribution and compared between groups with the appropriate parametric or non-parametric test. Most patients contributed a single recording (226 of 267 in the cohort), and Table 3 summarises patients, one value each. In the held-out set, 50 recordings came from 49 patients; clustering of recordings within patients was handled by keeping each patient’s recordings in one partition and by a patient-clustered bootstrap interval for the held-out AUC (Section 2.7). Separating a stable between-patient (trait) component from a within-patient (state) component would require repeated recordings across phases of illness, which this dataset does not contain (Section 3.2); that decomposition was therefore not performed.

Adjustment was pre-specified for platelet count, age and sex. Platelet count is included because it is the principal quantity already known to differ between the two phenotypes, and a difference that disappears once it is accounted for is a restatement of the platelet count rather than an independent observation. Age and sex are included as potential confounders of peripheral vascular reactivity. The role of each covariate and how it is handled are summarised in Table 2.

**Table 2.** Role of each covariate and how it is handled.

| **Variable** | **Role** | **Handling in the analysis** |
|---|---|---|
| **Platelet count** | Confounder; differs between phenotypes and partly defines them | Adjusted for (primary); also reported as a stand-alone reference |
| **Age and sex** | Potential confounders of peripheral vascular reactivity | Adjusted for |
| **Haematocrit rise, warning signs** | Mediators on the causal path from phenotype to signal | Not adjusted; used descriptively to define phase |
| **Body temperature** | Marker of illness stage and determinant of vasomotor tone | Sensitivity analysis |
| **Intravenous fluid** | Treatment acting directly on perfusion in the critical phase | Adjusted within the critical-phase stratum only (not recorded in the study cohort) |

**Table 3.** Characteristics of the finger-video cohort, the training partition and the held-out test set, by final diagnosis. Patient-level summaries except where stated. Laboratory values are the most extreme of the recorded visits, not of the whole admission.

|  | **Cohort — DF** | **Cohort — DHF** | **Training — DF** | **Training — DHF** | **Test set — DF** | **Test set — DHF** |
|---|---|---|---|---|---|---|
| Patients | 211 | 56 | 51 | 36 | 29 | 20 |
| Recordings | 249 | 62 | 58 | 42 | 30 | 20 |
| Recordings per patient, mean | 1.18 | 1.11 | 1.14 | 1.17 | 1.03 | 1.00 |
| Age, years, median (IQR) | 27 (21–40) | 36 (22–44) | 23 (19–36) | 36 (24–44) | 28 (21–39) | 32 (20–46) |
| Male, n (%) | 90 (43%) | 27 (48%) | 18 (35%) | 18 (50%) | 9 (31%) | 9 (45%) |
| Day of fever at first recording, median (IQR) | 5 (4–6) | 5 (5–6) | 6 (4–7) | 5 (5–6) | 5 (4–5) | 5 (5–6) |
| Lowest platelet count at recorded visits, × 10<sup>9</sup>/L, median (IQR) | 106 (82–130) | 38 (19–53) | 102 (77–115) | 36 (19–51) | 101 (72–122) | 39 (22–57) |
| Highest haematocrit at recorded visits, %, median (IQR) | 40 (36–42) | 42 (38–45) | 40 (37–42) | 42 (40–45) | 37 (35–41) | 39 (37–42) |

Not every variable that differs between the groups is a confounder, and treating them alike would be an error in the opposite direction. Haematocrit rise and the presence of clinical warning signs lie on the causal path between the phenotype and the optical signal: they are consequences of the leakage the phenotype produces, not competing explanations for it. Conditioning on a mediator removes the very effect the analysis is trying to estimate, so these variables are not adjusted for and are instead used descriptively to define phase. Body temperature occupies an intermediate position, being both a marker of illness stage and a direct determinant of peripheral vasomotor tone; it is therefore handled in a sensitivity analysis rather than in the primary model, with the reasoning stated rather than the choice made silently. The haematocrit level at the time of recording, as distinct from its rise, was examined descriptively, as a single-marker AUC (Mann–Whitney test; DeLong interval), by its correlation with the video probability (Spearman), and, as a post-hoc sensitivity analysis, by adding it to the platelet, age and sex model (Section 3.3). It was measured at the recording visit, not at its peak.

Intravenous fluid administration was pre-specified as a covariate to be adjusted for within the critical-phase stratum, because it is a treatment applied in one phase and acts on peripheral perfusion directly; it was not recorded in the study cohort, however, so this planned adjustment could not be performed (Section 3.3).

## 3. Results

### 3.1 Cohort

The finger-video cohort comprised 311 recordings from 267 patients: 249 recordings from 211 patients with a final diagnosis of DF and 62 recordings from 56 patients with a final diagnosis of DHF (Fig 2 and Table 3). Of the 267 patients, 226 contributed one recording, 39 contributed two, one contributed three and one four. No patient carried both diagnoses; four patients whose recordings were labelled inconsistently across visits had been removed before the cohort was frozen. Of the 62 DHF recordings, 58 were made while the patient was in the DHF state and 4 at recovery or discharge; DHF patients’ recordings from before leakage are not part of this dataset. Of the 249 DF recordings, 15 were made at recovery or discharge. DHF patients were slightly older than DF patients (median 36 versus 27 years, p = 0.14) and similar in sex (48% versus 43% male, p = 0.55).

After selection, 150 recordings from 136 patients (62 DHF from 56 patients; 88 DF from 80 patients) formed the modelling dataset: all 62 DHF recordings and 88 of the 249 DF recordings (Fig 2). Because DF recordings outnumbered DHF recordings four to one, the DF class was reduced to a size comparable with the DHF class to avoid a model dominated by one class, and all recordings were checked for comparable recording conditions. Selection was completed before any model was trained, without reference to any model output. The modelling dataset was split by patient into a training partition of 100 recordings from 87 patients (58 DF recordings from 51 patients; 42 DHF recordings from 36 patients) and a held-out test partition of 50 recordings from 49 patients (30 DF recordings from 29 patients; 20 DHF recordings from 20 patients); no patient contributed recordings to both (Fig 2). Within each diagnosis, the test partition was similar to the full cohort in median age, day of fever and lowest platelet count, although DF test patients were less often male (31% versus 43%) and haematocrit was slightly lower in both groups (Table 3). Within each diagnosis the training and test partitions were similar in age and sex (DF: age p = 0.42, sex p = 0.89; DHF: age p = 0.57, sex p = 0.94); in the training partition, DHF patients were older than DF patients (median 36 versus 23 years, p = 0.02) and, non-significantly, more often male (50% versus 35%, χ² p = 0.25; Table 3). These comparisons are descriptive. No test recording was excluded for signal quality.

### 3.2 Phenotype separation on the held-out recordings

All held-out DHF recordings were obtained at or after leakage onset. This model therefore estimates separation of the observed DF and DHF states; it cannot estimate a phenotype-by-phase interaction. Expectation (i), separation at least as good as platelet count, was assessed by the held-out AUC and the paired comparison on recordings with a platelet measurement (Section 3.3).

Eight network architectures, covering convolutional, residual, recurrent and attention-based designs, were trained under the identical protocol described in Section 2.7 and S1 Text and compared on the held-out test set. DeepPhys-1D was retained as the final model under the pre-specified ranking rule (Section 2.7); held-out AUCs of all eight architectures ranged from 0.860 to 0.963 (S1 Fig).

On the 50 held-out recordings the final model separated DHF from DF with an AUC of 0.963 (DeLong 95% CI 0.918– 1.000; patient-clustered bootstrap 0.906–0.998; permutation p < 0.0001). At the pre-specified threshold of 0.50 it classified 45 of 50 recordings correctly: 18 of 20 DHF recordings (sensitivity 90.0%, 95% CI 69.9–97.2%) and 27 of 30 DF recordings (specificity 90.0%, 74.4–96.5%) (Fig 3, Table 4). The accuracy of 90.0% (78.6–95.7%) compares with 60% for a rule that calls every recording DF, and the balanced accuracy was 0.900. The predictive values, 85.7% and 93.1%, apply to the test-set prevalence of 40% DHF, twice the 20% of the cohort, and would be different in any other population.

**Fig 3.** Classification of the 50 held-out recordings at the pre-specified threshold of 0.50. DHF is the positive class. (A) Confusion matrix against the final clinical diagnosis, with row and column totals. (B) Sensitivity, specificity, predictive values and accuracy with 95% Wilson intervals; the vertical mark shows the 60% accuracy obtained by calling every recording DF. Predictive values depend on the 40% DHF prevalence of the test set.

**Table 4.** Performance of the final model on the held-out test set (50 recordings; 20 DHF, 30 DF; DHF positive). Threshold 0.50, fixed in advance.

| **Measure** | **Estimate** | **95% CI** | **Counts** |
|---|---|---|---|
| AUC | 0.963 | 0.918–1.000 (DeLong); 0.906–0.998 (patient bootstrap) | 20 DHF, 30 DF |
| Average precision (area under precision–recall curve) | 0.957 | — | no-skill value 0.40 |
| Sensitivity (DHF called DHF) | 90.0% | 69.9–97.2% | 18/20 |
| Specificity (DF called DF) | 90.0% | 74.4–96.5% | 27/30 |
| Positive predictive value | 85.7% | 65.4–95.0% | 18/21 |
| Negative predictive value | 93.1% | 78.0–98.1% | 27/29 |
| Accuracy | 90.0% | 78.6–95.7% | 45/50; majority-class rate 60% |
| Balanced accuracy | 0.900 | — | — |
| Brier score | 0.087 | — | prevalence-only reference 0.240 |
| Calibration-in-the-large (ideal 0) | −0.37 | −1.10 to 0.36 | mean predicted 0.455; observed 0.400 |
| Calibration slope (ideal 1) | 2.59 | 0.85–4.33 | — |
| Selection-free reference: mean of all 8 architectures, AUC | 0.957 | 0.909–1.000 | 20 DHF, 30 DF |
| Clinical reference: platelet count alone, AUC | 0.888 | 0.795–0.980 | 19 DHF, 30 DF |

The five errors were two DHF recordings with probabilities of 0.30 and 0.46 and three DF recordings with probabilities of 0.53, 0.58 and 0.61; four of the five lay within 0.15 of the threshold, and in all five the finger was detected on every frame, so none arose from a failure to find the finger (Fig 4C). The mean probability assigned to the predicted class was 0.80 for correct and 0.59 for incorrect decisions (0.41 for the correct class in the incorrect ones).

**Fig 4.** Discrimination and calibration on the held-out recordings. (A) ROC curve of the video model (red) and of platelet count alone on the same recordings (grey, dashed; one DHF recording lacked a platelet count). The open circle marks the 0.50 threshold. (B) Observed proportion of DHF against mean predicted probability in quintiles of predicted probability, with 95% Wilson intervals; the diagonal is perfect calibration. (C) Distribution of predicted probability in bins of 0.05, DHF recordings above the axis and DF recordings below; bars on the wrong side of the dashed threshold are the five errors.

Selection among architectures contributed little. The eight candidate architectures reached AUCs of 0.860–0.963 (median 0.946; S1 Fig); the selected model did not differ significantly from the second-ranked (difference 0.020, 95% CI −0.039 to 0.079, p = 0.50); and the unweighted average of all eight architectures, which involves no selection on the test set, reached 0.957 (0.909–1.000).

Calibration was adequate in the middle of the range but too conservative at the extremes (Fig 4B). The Brier score was 0.087, against 0.240 for a model that predicts the prevalence for everyone. Calibration-in-the-large was −0.37 (−1.10 to 0.36), consistent with no overall bias, but the calibration slope was 2.59 (0.85–4.33): the probabilities appear compressed towards 0.5 (the interval for the slope includes 1), so that no DHF recording occurred among the 20 with the lowest probabilities (mean predicted 0.14) and all 10 with the highest probabilities (mean 0.91) were DHF. In practical terms, the scores distinguish the observed groups well, but the probabilities are compressed towards 0.5. They should not be interpreted as calibrated absolute risks without recalibration and validation in the intended population.

The broader phenotype-by-phase and phase-specific questions cannot be estimated from this model. All 20 DHF test recordings, and 58 of the 62 DHF recordings in the cohort, were made while the patient was in the DHF state, and the dataset contains no pre-leakage DHF recording; by day of fever the DHF test recordings were from day 4 (1), day 5 (12), day 6 (4) and days 7–9 (3). The result in the study cohort is therefore a comparison between DHF recorded at or after the onset of leakage and DF, and it cannot distinguish a stable property of the DHF patient from a property of the leaking state. Probability did not track day of fever within either group (Spearman ρ 0.28, p = 0.13, in DF; 0.15, p = 0.52, in DHF). The clinically important comparison of pre-leakage DHF against DF requires a separate model evaluated on those groups.

### 3.3 Adjustment for platelet count and other covariates

Expectations (ii)–(iv), concerning information beyond platelet count, were evaluated through added-value modelling, the comparison with thrombocytopenic DF and within-diagnosis correlations. The direct AUC comparison first completes the assessment of expectation (i).

In the held-out recordings the platelet count was a median of 39 × 10⁹/L (IQR 22–57) in DHF and 102 × 10⁹/L (73– 122) in DF; 18 of 19 DHF recordings with a count (95%) and 14 of 30 DF recordings (47%) were at or below 100 × 10⁹/L. Alone, platelet count separated the groups with an AUC of 0.888 (0.795–0.980) in the same 49 recordings, against 0.961 for the video; the difference, 0.074 (−0.036 to 0.184, p = 0.19), was not statistically significant in a set of this size (Table 5, Fig 4A). The lower confidence limit excludes a deficit of the video of more than 0.036 in AUC, consistent with expectation (i).

**Table 5.** Adjustment of the video signal for platelet count, age and sex. All analyses use the held-out recordings and are exploratory.

| Analysis (held-out recordings with a platelet count; 19 DHF, 30 DF) | Estimate (95% CI) | Test |
|---|---|---|
| **Direct comparison on the same recordings** |  |  |
| Platelet count alone, AUC | 0.888 (0.795–0.980) | — |
| Haematocrit alone, AUC | 0.581 (0.409–0.753) | Mann–Whitney p = 0.35; 49 recordings (19 DHF, 30 DF) |
| Video probability alone, AUC | 0.961 (0.914–1.000) | difference 0.074 (−0.036 to 0.184), p = 0.19 |
| DHF against DF with platelets ≤ 100 × 10<sup>9</sup>/L (19 DHF, 14 DF): video AUC | 0.970 (0.922–1.000) | platelets in the same subset 0.759 (0.582–0.936); difference 0.211 (0.020 to 0.401), p = 0.03 (paired DeLong) |
| **Added value: leave-one-out cross-validated AUC** |  |  |
| Platelet count | 0.865 | — |
| Platelet count + age + sex | 0.846 | — |
| Platelet count + age + sex + haematocrit † | 0.844 | — |
| Video | 0.958 | — |
| Video + platelet count | 0.981 | vs platelet count: +0.116 (0.027–0.204), p = 0.010; likelihood-ratio χ<sup>2</sup> = 33.6, p < 0.001 |
| Video + platelet count + age + sex | 0.977 | vs platelets + age + sex: +0.132 (0.039–0.224), p = 0.005; likelihood-ratio χ<sup>2</sup> = 31.4, p < 0.001 |
| Video + platelet count + age + sex + haematocrit † | 0.977 | vs platelets + age + sex + haematocrit: +0.133 (0.039–0.228), p = 0.006; likelihood-ratio χ<sup>2</sup> = 31.0, p < 0.001 |

† Post-hoc sensitivity analysis; haematocrit was measured at the recording visit, not at its peak.

Three analyses asked whether the video carries information the platelet count does not (Fig 5). First (iv), within each diagnosis the video probability was unrelated to the platelet count (Spearman ρ −0.18, p = 0.35, in DF; 0.05, p = 0.84, in DHF); the correlation across all recordings (ρ −0.53) reflects the difference between the groups rather than a dependence of one measure on the other. Second (iii), among the recordings that platelet count cannot readily separate — DHF against thrombocytopenic DF with counts of 100 × 10⁹/L or less — the video retained an AUC of 0.970 (0.922–1.000), whereas platelet count fell to 0.759 (0.582–0.936); in this exploratory comparison the paired difference was 0.211 (0.020 to 0.401, p = 0.03; paired DeLong), consistent with expectation (iii): once DF was restricted to low platelet counts, platelet count lost most of its advantage and the video did not. The lower confidence limit is close to zero and the comparison was not adjusted for multiple testing. Third (ii), adding the video to a model containing platelet count improved fit (likelihood-ratio χ² = 33.6, p < 0.001) and cross-validated separation (AUC 0.865 to 0.981; difference 0.116, 0.027–0.204, p = 0.010), and the result was unchanged after adding age and sex (0.846 to 0.977; p = 0.005). Because the video score separated the groups almost completely, odds ratios could not be estimated reliably in 49 recordings and are not reported; the added value of the video is shown by the likelihood-ratio test and cross-validated AUC.

**Fig 5.** Does the video signal survive the platelet count? (A) Video probability against platelet count for each held-out recording with a count; the horizontal line is the 0.50 threshold and the dashed line 100 × 10⁹/L. (B) Leave-one-out cross-validated AUC of logistic models built from platelet count, age, sex and the video probability, with DeLong 95% intervals. (C) AUC of the video model in subsets and sensitivity analyses (red; the one-recording-per-patient analysis was post hoc), with platelet count in the thrombocytopenic subset for comparison (grey).

A raised haematocrit is part of the leakage process, so we asked whether the video could simply be reflecting it. Haematocrit measured at the time of recording did not separate the held-out groups: the median was 39% (IQR 37–42) in DHF and 37% (35–41) in DF, and the AUC was 0.58 (DeLong 95% CI 0.41–0.75; Mann–Whitney p = 0.35) in the 49 recordings with a value, against 0.961 for the video (Table 5). Among the recordings with platelet counts of 100 × 10⁹/L or less it was 0.57 (0.36–0.77). The video probability was unrelated to haematocrit (Spearman ρ 0.00, p = 0.98 across all recordings; −0.25, p = 0.30 within DHF; −0.07, p = 0.71 within DF). As a post-hoc sensitivity analysis, haematocrit was added to the platelet, age and sex model; it contributed nothing (likelihood-ratio χ² = 1.55, p = 0.21; cross-validated AUC 0.846 to 0.844), and the video still added to that model (AUC 0.977 against 0.844; difference 0.133, 0.039–0.228, p = 0.006; likelihood-ratio χ² = 31.0, p < 0.001; Table 5). In the full modelling set of 150 recordings (145 with a value), DHF recordings had a modestly higher haematocrit (median 42%, IQR 38– 45, against 39%, 36–42; AUC 0.63, 0.54–0.73; p = 0.007), a small difference that is far short of what the video achieved. Haematocrit was measured at the recording visit rather than at its peak, and may reflect fluid already given, so these results do not exclude a relation at other times.

Age and sex were not model inputs, but in the training partition DHF patients were older than DF patients (median 36 versus 23 years, Mann–Whitney p = 0.02; Table 3), so we checked, post hoc, whether the model had learned demographic correlates. In the held-out recordings, age and sex alone did not separate the groups (AUC 0.55 and 0.58; DeLong 95% CI 0.37–0.73 and 0.44–0.72); the video probability was unrelated to age within either diagnosis (Spearman ρ −0.15 in DF, p = 0.43; −0.17 in DHF, p = 0.48) and did not differ between men and women (Mann– Whitney p = 0.89 in DF, 0.45 in DHF); and discrimination was similar in women and men (AUC 0.991 and 0.926; p = 0.27) and in patients younger and older than 30 years (0.967 and 0.953; p = 0.79). These subgroups are small.

Only 6 of the 50 held-out recordings were made at a temperature of 37.5 °C or above (5 DF, 1 DHF). Temperature did not separate the groups (AUC 0.44) and was unrelated to the video probability (ρ −0.06, p = 0.69). Restricted to the 44 afebrile recordings, the AUC was 0.968 (0.925–1.000). The study cohort therefore contains too few febrile recordings to test the effect of fever itself.

The bedside pulse rate did not separate the groups (AUC 0.53) and was unrelated to the video probability (ρ 0.01). Performance was unchanged when restricted to one recording per patient (AUC 0.962) (Fig 5C).

Differences between videos in lighting and in the size of the finger in the frame can be seen by eye, and a difference in recording conditions between the groups could in principle produce a difference in the signal that has nothing to do with the patient. Recording conditions were therefore controlled rather than assumed to be equal. All recordings were made with the same phone under the same protocol; all recordings were checked for comparable lighting and fingertip size in the frame; and the analysis removes overall brightness and colour balance from every signal (Sections 2.5 and 2.6). In addition, the model never sees the image itself, only the fluctuation of fingertip colour over time, so background, framing and skin tone do not enter its input.

Haematocrit rise and the warning signs were treated as intermediates on the causal pathway from phenotype to signal and were not adjusted for in the primary analyses (Section 2.8); haematocrit level was examined only as a post-hoc sensitivity analysis (Section 3.3). Intravenous fluid administration was pre-specified as a covariate (Section 2.8) but is not recorded in the study cohort and could not be examined.

## 4. Discussion

In our study, fingertip video was able to differentiate dengue haemorrhagic fever (DHF) in plasma leakage from dengue fever (DF) with high discrimination in this internal test set (AUC 0.963; 0.957 for the unweighted average of eight candidate architectures, which involves no selection on the test set). On the 49 recordings with a platelet count, the video-derived probability (AUC 0.961) was not clearly different from contemporaneous platelet count (AUC 0.888): it was numerically higher, but the paired difference did not reach statistical significance. The rPPG signals retained strong separation in the clinically decisive comparison of DHF against thrombocytopenic DF (AUC 0.970 versus 0.759 for platelets; exploratory paired p = 0.03) and, in exploratory analyses, improved model fit and cross-validated discrimination when added to platelet count. These findings suggest that a short optical recording of the fingertip carries information complementary to the platelet count, within the post-onset window and single-centre internal test set studied here; they do not, however, establish early prediction, statistical independence in a general population, or clinical utility yet.

Dengue fever is associated with systemic viral inflammation, while dengue haemorrhagic fever results in an amplified immune response with higher levels of immune complex formation, complement activation and massive cytokine release [38,39]. The main pathophysiological factor distinguishing DHF from DF is plasma leakage associated with increased vascular permeability [39]. Glycocalyx disruption increases vascular permeability in dengue [40]. Vascular hyperpermeability ultimately leads to hypovolaemia and extravascular fluid accumulation with dengue shock syndrome. The available treatment is supportive fluid management. It is important to identify patients at risk of DHF as early and accurately as possible. Current diagnostic practice relies on monitoring platelet count, clinical red flags, and detecting a rise in haematocrit of more than 20% above baseline. This relies on full blood counts or fingertip/bedside haematocrit readings, with support from bedside ultrasonography, which has been shown to detect plasma leakage before it crosses the conventional haemoconcentration threshold in a proportion of cases [41].

Whether an optical fingertip signal shares this early sensitivity is untested in the present study; Section 1.3 sets out three physiological routes — endothelial, haemodynamic and optical — by which a leakage state could plausibly alter such a signal, and this section returns below to what the present results can and cannot say about which route, if any, operates.

In a prospective Vietnamese study of 250 hospitalised adults and children with dengue, an artificial-intelligence model applied to 24-hour wearable PPG classified volume status, defined from the clinical timing of shock, with a sensitivity of 0.81 and a specificity of 0.70 [8].

Two further Vietnamese cohorts used continuous contact PPG from wearable sensors: a multimodal transformer forecast clinical state 2 hours ahead in 153 patients, identifying low-risk states (NEWS2 and mSOFA < 6) with an AUROC of 0.83 and an AUPRC of 0.67 [9], and a convolutional neural network classified 132 adults into three severity classes (precision 0.79, recall 0.77) and 15 children in the acute versus convalescent phase (precision and recall 0.89) under internal cross-validation [10]. All three studies were conducted in Vietnam, a country with a high dengue burden like Sri Lanka. Our study differs from them in using a single short, non-contact smartphone fingertip video rather than continuous monitoring with a contact wearable. The question also differs. In those studies the labels were operational: the clinical timing of shock [8], composite early-warning scores [9], or the location of care (ward, emergency department or intensive care unit) [10], and performance was judged by how closely a model reproduced them. Here the label was the pathophysiological distinction itself, plasma leakage in DHF against its absence in DF (Section 2.3), and the video signal was evaluated as a candidate marker of that process rather than as a classifier alone. It was set against the existing reference marker, the platelet count, through four expectations that could each have failed (Sections 1.4 and 1.5): comparable separation, added information, retained separation where platelet counts overlapped, and no correlation with the count within either group. The held-out results were consistent with all four (Section 3.3), although these analyses were exploratory and internal.

Smartphone fingertip video has previously been used to estimate platelet count [11]; the present signal does not appear to reproduce that estimate, because it was unrelated to the platelet count within each diagnosis (Section 3.3). In this test set the video score was not explained by heart rate, temperature or haematocrit, although the absence of a detected correlation in 49 recordings does not exclude a relation. However, the current study cannot attribute the signal to a specific physiological mechanism among the several plausible routes discussed in Section 1.3, since no feature-attribution or channel-removal analysis was performed; the model’s output is interpreted only as a whole.

There are several other limitations in our study. All results come from a single centre, one device and one standardised recording protocol, and the held-out set is small and was drawn from the same cohort by patient-level partition; they are therefore internal validation only, and performance with other operators, devices, recording conditions or centres may be lower. External validation in independent, prospectively collected cohorts at other sites is required before any conclusion about generalisability or clinical use can be drawn. The optic signals remain sensitive to lighting, motion, and hardware settings. The model probability outputs were poorly calibrated at extremes (Brier score 0.087; calibration slope 2.59, suggesting probabilities compressed toward 0.5) and should not be read as absolute clinical risk scores. Intravenous fluid administration, which can itself alter peripheral perfusion, was not recorded in this dataset and could not be examined as a potential confounder; both DF and DHF patients received a combination of intravenous and oral fluids as part of routine care, so differential fluid treatment is not expected to explain the observed separation, but it cannot be formally excluded. The DF class is a subset of 88 of 249 recordings chosen for fingertip geometry similar to the DHF recordings, and the test set is 40% DHF against 20% in the cohort, so specificity and predictive values apply to that subset and prevalence and may differ in unselected DF. The held-out test set was also used once to select among eight candidate architectures, so the reported performance of the chosen model is likely somewhat optimistic; the eight-architecture average (AUC 0.957) is a more conservative estimate. The added-value estimates in Section 3.3 use scores from the same selected model and therefore share this optimism. Haematocrit was measured at the recording visit rather than at its peak and may reflect fluid already given, and its handling as a post-hoc covariate departs from its pre-specified role as a mediator. Day of fever was patient-reported and subject to recall error, which may reduce the precision of any fever-day-specific comparisons. We used a convolutional neural network approach; other approaches, including random forests and principal component analysis, are yet to be explored. The model was trained on video-derived signals only; age, sex and other clinical variables were not inputs. Because DHF patients in the training partition were older than DF patients (median 36 versus 23 years) and, non-significantly, more often male (50% versus 35%), the network could still have learned demographic correlates of the signal implicitly. We therefore examined performance by sex and age band and the relation of the video score to age (Section 3.3): discrimination was similar across these subgroups, the video score was unrelated to age, and the video signal retained its added value after adjustment for age, sex and platelet count (Table 5). The subgroups were small, and the imbalance remains a limitation that should be addressed by demographic matching in external validation.

Our findings suggest that a finger rPPG colour signal separates DHF recorded in plasma leakage from DF, and that, in exploratory analyses of this test set, this separation was not explained by platelet count. This study is a proof of concept for an association between fingertip rPPG signals and the leaking state; it does not show that the signal reflects microvascular changes, or that it can detect leakage before it begins. Whether it can do so is the question for future pre-leakage studies.

With this proof of concept, we would like to present recommendations to future research. The comparison markers should be widened. The video signal should be compared against serial haematocrit rise and ultrasonographic leakage detection. Pre-leakage testing should be performed to evaluate whether rPPG can detect the onset of plasma leakage at least a few hours earlier. If so it can be used for early risk prediction. External validation should be done across different devices, lighting environments, skin tones, and clinical centres. Most importantly, the exact physiological mechanism should be validated — using endothelial biomarkers to test the endothelial route, and a simple postural (seated-to-standing) provocation to test the haemodynamic route directly.

## Data Availability

The video recordings and identifiable clinical data cannot be shared publicly, as participants did not consent to public release of the recordings this restriction was imposed by the Ethics Review Committee of the National Institute of Infectious Diseases (NIID), Sri Lanka (ERC Reference No. P/04/02/2026). De-identified per-recording predictions are provided in S1 Data. Requests for access to further de-identified data may be addressed to the Ethics Review Committee, NIID, Sri Lanka. Code: The source code and trained model are proprietary and are not publicly released. Editors and reviewers may request confidential access to the code and trained model through the engineering team at Hii.Health Pte. Ltd. other groups can submit recordings for evaluation under a confidentiality agreement.

## Data availability statement

The video recordings and identifiable clinical data cannot be shared publicly, as participants did not consent to public release of the recordings; this restriction was imposed by the Ethics Review Committee of the National Institute of Infectious Diseases (NIID), Sri Lanka (ERC Reference No. P/04/02/2026). De-identified per-recording predictions are provided in S1 Data. Requests for access to further de-identified data may be addressed to the Ethics Review Committee, NIID, Sri Lanka.

## Code availability

The source code and trained model are proprietary and are not publicly released. Editors and reviewers may request confidential access to the code and trained model through the engineering team at Hii.Health Pte. Ltd. The trained model can also be evaluated under a confidentiality agreement: other groups can submit recordings and receive the model’s predictions. The per-recording probabilities for the held-out test set are provided (S1 Data), so that the model’s held-out performance (AUC, sensitivity, specificity, predictive values and calibration) can be recomputed; analyses that also use clinical variables (platelet count, haematocrit, age and sex) require clinical data that are not included in S1 Data, and the same processing code is used during training and in the deployed software. Every processing step is specified in Section 2.6 and S1 Text, using published algorithms [23,24,33], so that an equivalent model could be rebuilt from the description.

## Supporting information

**S1 Text. Technical detail of signal extraction and model development.** The full processing specification for fingertip localisation, signal extraction, the signal-quality rule, model architecture and training, and the eight candidate architectures (file S1_Text.docx).

**S1 Fig. Held-out performance of the eight candidate architectures** (file S1_Fig.tif), trained under the identical protocol described in S1 Text. Left: AUC with DeLong 95% intervals (trainable parameters per network in brackets). Right: specificity (circles) and sensitivity (squares) at the 0.50 threshold, with balanced accuracy. DeepPhys-1D was selected by the rule in Section 2.7. Architectures: DeepPhys-1D, CNN-LSTM (convolutional front end with bidirectional recurrent layers), ResNet-1D, InceptionTime, CNN-1D, PhysFormer-1D, CyFormer-1D (hybrid convolution–transformer) and a transformer encoder.

**S2 Fig. Signal construction for one held-out DHF recording** (file S2_Fig.tif). From top: mean red, green and blue inside the fingertip region (grey, trimmed segments); green trace resampled to 30 Hz with its 1.5-s baseline; green expressed relative to baseline before and after 0.5–4 Hz filtering; POS and CHROM pulse signals; one 10-s classifier window.

**S1 Checklist. TRIPOD+AI reporting checklist**, completed for this study, with the section reporting each item (file S1_Checklist.docx).

**S1 Data. Final diagnosis and predicted probability of DHF for each of the 50 held-out test recordings**, with patient identifiers removed (file S1_Data.csv).

